# Validation of an electronic-nose sensor array in detecting *C. difficile* and differentiating toxin-positive from toxin-negative stool samples

**DOI:** 10.64898/2026.10.06.26364614

**Authors:** Patrick N. Balius, Christopher Campbell, Muhammad Ikrma, Hien H. Nguyen

## Abstract

**Objective:** To evaluate the specimen-level performance of a multichannel electronic nose (e-nose) sensor array in classifying stool specimens into relevant *Clostridioides difficile* categories.

**Design:** Retrospective single-center validation study with reference to polymerase chain reaction (PCR) and *C. difficile* toxin testing.

**Participants:** A convenience sample of 31 discarded stool specimens with PCR and confirmatory toxin testing. Categorized into PCR-positive (+)/toxin+, PCR+/toxin-negative (–), and PCR–.

**Methods:** Specimens were sampled in controlled headspace by two e-nose sensor arrays in 10-minute sessions with 5-second sampling intervals. The primary endpoint was one-versus rest classification performance (sensitivity, specificity, positive predictive value, and accuracy) into three classes: PCR+/toxin+, PCR+/toxin–, and PCR–. Data were normalized and the region of interest was extracted for training and testing. Random forests were utilized for classification of normalized data with a test-train split at the specimen-level.

**Results:** There were 457 unique 10-minute sampling events (147 PCR+/toxin+, 183 PCR+/toxin–, 127 PCR–). Sensors 1 and 2 captured 20,331 and 23,538 unique 5-second exposures within the region of interest, respectively. Testing data classification accuracy was 0.79 [0.78, 0.81] and 0.84 [0.83, 0.85] for Sensors 1 and 2. Sensitivity for PCR+/toxin+ exposures was 0.82 [0.79, 0.84] and 0.94 [0.92, 0.95], and specificity was 0.91[0.90, 0.93] and 0.93 [0.92, 0.94] for Sensors 1 and 2.

**Conclusion:** An e-nose sensor array partially differentiated PCR-classified *C. difficile* specimen classes at the exposure level under headspace-controlled conditions. Performance was strongest for PCR+/toxin+ exposures. Findings support further prospective validation in the hospital room setting.

## Introduction

*Clostridioides difficile* infection (CDI) is the leading cause of healthcare-associated diarrhea in the United States, with an estimated 183,000 healthcare-associated infections (HAIs) per year.^1^ Hospital-onset CDI is estimated at 8.3 cases per 10,000 patient days, resulting in an additional 3 to 21 days in length of stay compared to non-CDI patients.^2^ Incidence and severity increases with age, as complications like toxic megacolon, pseudomembranous colitis, and sepsis are potentially life-threatening in higher-risk populations; those 65 and older have an in-hospital mortality rate of 8.5%, nearly 15 times the rate of young adults.^3,4^ This results in a general strain on healthcare system resources including direct and indirect costs, staffing, and potential outbreaks.

Diagnosis currently relies primarily on five testing methods, either in combination or as stand-alone methods. The gold standard references are toxigenic culture (TC) and cytotoxicity neutralization assay (CNNA), whereas the most frequently used methods are enzyme immunoassays (EIAs) of toxins A and B, EIAs of glutamate dehydrogenase (GDH), and nucleic acid amplification tests (NAATs), usually via polymerase chain reaction (PCR). NAATs have the highest sensitivity among rapid tests, but have lower specificity than toxin EIAs because, unlike toxin EIAs, NAATs test for the presence of the toxins A and B genes, not the presence of the toxins themselves; this risks the detection of asymptomatic colonization by toxigenic *C. difficile.*^5^ The Infectious Disease Society of America recommends a two-step algorithm based on a sensitive first-step test (NAAT or EIA for GDH) in combination with a more specific test (EIA for toxins A and B or CNNA) to rule out *C. difficile* colonization without the presence of free toxins.^6^ Although effective for point-of-care diagnosis, these methods provide point-in-time confirmation and do not enable continuous environmental or non-contact detection.

Electronic nose (e-nose) devices present a potential solution for continuous monitoring and detection with more timely resource-and-cost-saving interventions than the standard of care tests alone. E-noses are devices that mimic mammalian olfaction by detecting complex volatile organic compound (VOC) patterns—the volatilome—via multichannel sensor arrays and analyzing the volatilome via various algorithmic classification methods.

Due to a unique VOC profile from sulfur-producing amino acids integral to the bacteria’s metabolism, *C. difficile* is a good candidate for e-nose detection. This application has been studied priorly to detect *C. difficile* in both stool specimens and patient breath samples via e-nose.^7,8^ The current study aimed to evaluate the ability and performance of two unique e-nose sensor arrays in differentiating between three *C. difficile* classes of stool specimens with reference to prior testing and confirmation via PCR and toxin testing: PCR-positive (+) for *C. difficile* and toxin+, PCR+ for *C. difficile* and toxin-negative (–), and PCR– for *C. difficile*. The e-nose utilized in this study employs an array of chemically sensitive carbon nanotube-based nanosensors with varying reactivity to various VOCs; the carbon nanotube sensors utilized are based on previous development of nanotube formulations for the detection of COVID-19 in human breath.^9^ This was done within a clinical laboratory environment.

## Methods

### Study Design

This pre-screening tool validation study was conducted at a Veterans Affairs (VA) Medical Center with a large clinical research laboratory. The study was approved by the Northern California VA IRB. A convenience sample of discarded fecal specimens processed at the VA Medical Center was collected for this study. Fecal specimens that had been tested via PCR for *C. difficile* were eligible for inclusion if the samples were PCR– for *C. difficile* or PCR-positive for *C. difficile* with confirmatory EIA toxin results. Eligible specimens were de-identified and categorized into three classifications by PCR and toxin EIA reference result: PCR+/toxin+, PCR+/toxin–, and PCR–.

### Sample Handling and Exposure

Specimens were maintained in their original sample storage containers. Containers were equilibrated to room temperature and gently mixed for greater VOC availability prior to testing. The experimental setup contained a sampling chamber with two e-nose devices (hereafter Sensor 1 and Sensor 2). The sampling chamber was connected in parallel to a circulatory system with a rheostat-controlled gas pump. For each stool specimen sampling event, the sampling chamber was first flushed for 10 minutes with synthetic air pumped through an active charcoal scrubber to establish a baseline for later normalization. Stool specimens were then introduced to the circulatory system and concomitant headspace gases were introduced via gas pump to the sampling chamber and allowed to circulate for 10 minutes. The sampling chamber was then off-gassed for 5 minutes before repeating the chamber scrubbing and sampling protocol. Each e-nose sensor had a sampling rate of once every five seconds, resulting in time-series data that comprised the instantaneous response of all 16 sensor channels at each five-second measurement.

### Algorithmic Testing and Training and Statistical Analysis

Following specimen sampling by the e-nose sensors, the mean baseline response for each channel was computed from the ambient measurements immediately preceding gas exposure.

Sensor response data were normalized relative to the synthetic air baseline in order to remove sensor drift and allow comparability across exposures and sensors. This was done by converting raw sensor resistance changes into percentage differences from ambient conditions. The normalized response for each sensor channel, the Electrical Parameter Value (EPV), was calculated as:

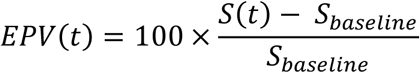

where *t* is a given time instant, measured at five second intervals, at which sensor telemetry data was recorded; *S(t)* represents the raw sensor signal at time *t*; *S_baseline_* denotes the mean baseline signal measured during the ambient air phase immediately prior to exposure; and *EPV(t)* represents the percentage change in sensor response relative to baseline.

After normalization, the region of interest (ROI) corresponding to the active response to VOCs was isolated and extracted from the e-nose temporal signal. The ROI was identified using a trend-based detection approach. The beginning of the ROI was defined as the first index where a sustained positive derivative sequence occurred, indicating a consistent upward response of the sensor signal for five consecutive samples:

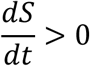

The end of the ROI was defined as the first index in which a sustained negative derivative sequence occurred, indicating a consistent downward response of the sensor signal for five consecutive samples. The ROI therefore spans the interval between the onset of the sensor response and the point where the signal approaches the baseline reference level. The resulting ROI contains all time points corresponding to the gas-sensor interaction phase, which forms the basis for subsequent feature extraction and model training. An example of the ROI within a given sampling event is shown in Figure 1.

**Figure 1.**
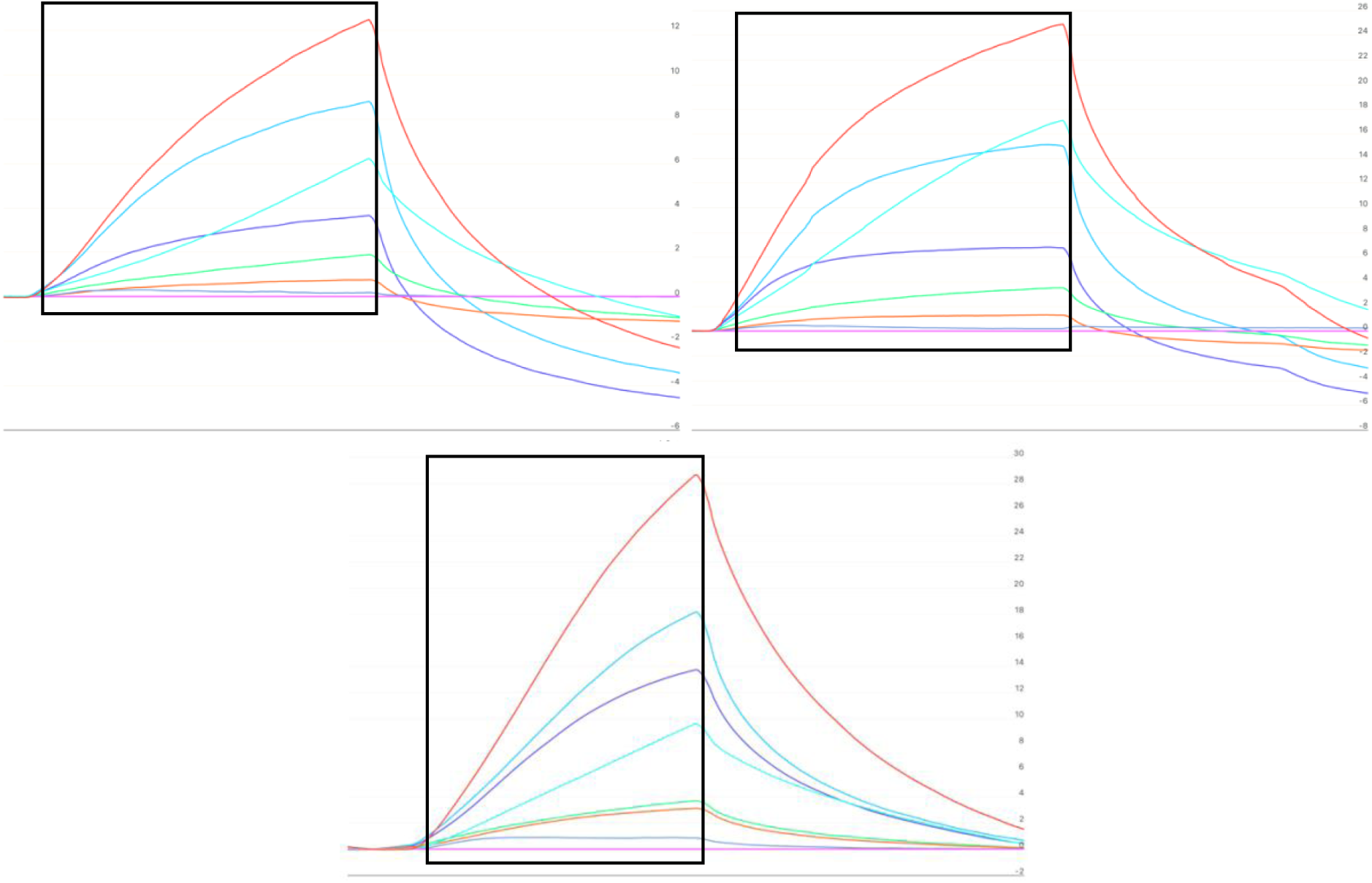
Example of sensor response curves during e-nose sampling events for PCR-positive/toxin-positive (top), PCR-positive/toxin-negative (middle), and PCR-negative (bottom) stool specimens. Each line corresponds to an ink solution, each of which makes up two sensors within the sensor array. The section within the rectangle represents the region of interest (ROI) extracted for exploratory analysis and classification modeling.

### Exploratory Analysis and Model Selection

Following normalization and ROI extraction, the sensor array responses were structured into a feature representation suitable for machine learning analysis. The e-nose sensor system records time-series responses from a 16-channel array, where each measurement corresponds to the instantaneous response of all sensor channels. For each sensor channel *i* at time step *t*, the sensor array response was represented as a feature vector composed of the normalized EPVs from all 16 channels where *EPV_i_(t)* denotes the normalized percentage response of sensor channel *i* at time *t* for channels 1 through 16:

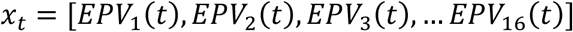

Each vector represents the multidimensional response pattern of the sensor array at a given measurement point during a sampling event. All observations across sampling events collectively form the feature matrix:

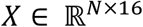

where N denotes the total number of observations (i.e., every time point in the ROI) and 16 corresponds to the number of sensor channels.

To examine the separability of the study classes within this feature space, Linear Discriminant Analysis (LDA) was applied as an exploratory dimensionality reduction technique. LDA identifies projection directions that maximize separation between class means relative to the variability within each class. Projection of the normalized sensor responses into the LDA space enabled the visualization of the class distribution within the feature space and provided an initial assessment of class separability prior to model training. The LDA visualization indicated a separable structure within the normalized sensor response data.

Given evidence of class separability, seven machine learning models, representing distinct algorithmic paradigms, were trained and compared to comprehensively evaluate classification performance. These included logistic regression, support vector machine, k-nearest neighbors, decision tree, random forest, extra trees, and XGBoost. This selection ensures broad methodological coverage across linear, distance-based, boundary-based, and ensemble approaches to classification. Each model was trained to classify observations into the three study classes using the normalized sensor response vectors—hereafter referred to as “exposures”—as input features. Model selection was based on overall test accuracy and sensitivity for the PCR+/toxin+ class, as correct identification of toxin-positive *C. difficile* infection represents the primary clinical objective of this diagnostic tool.

Prior to model development and evaluation, the dataset was partitioned into training and testing sets to evaluate model generalization with splits between 79-85% and 15-21%, respectively. The split was performed at the sampling event level, such that all observations derived from a single sampling event were assigned entirely to either the training set or the testing set. This was applied across all study classes to ensure that exposures from each class were represented in both training and testing partitions. The result was two independent datasets for model performance evaluation, one for training and one for testing.

### Performance Evaluation Metrics

Classification performance was evaluated for each sensor array at the exposure level using one-versus-rest methodology (e.g., differentiation of the PCR+/toxin+ reference class from the PCR+/toxin– and PCR– classes combined).^10^ Calculated performance metrics include sensitivity, specificity, accuracy, and positive predictive value. Due to the three-class classification, each reference result class—including the negative class—had these metrics calculated. Wilson score confidence intervals with a 95% confidence level were calculated for each performance metric. As multiple exposures were derived from individual specimens, observations are clustered and not statistically independent. Confidence intervals are therefore interpreted descriptively and may underestimate variance due to within-specimen correlation.

## Results

The cohort consisted of 31 specimens: 11 PCR+/toxin+, 10 PCR+/toxin–, and 10 PCR–. There were a total of 147 PCR+/toxin+, 183 PCR+/toxin–, and 127 PCR– sampling events. Based on accuracy and sensitivity for the PCR+/toxin+, random forest models were chosen for the classification of data from Sensors 1 and 2. Model comparisons can be found in the supplementary material in Tables S-1 and S-2.

### Sensor 1

A total of 20,331 (6,893 PCR+/toxin+, 8,877 PCR+/toxin–, and 4,561 PCR–) 5-second exposures were captured by Sensor 1. These were split into training and testing datasets, further detailed in Table 1. LDA plots revealed that each class formed clusters with some overlap between classes, indicating partial class separability (Figure 2). For testing data, overall accuracy was 0.79 (95% CI: 0.78, 0.81) with high discrimination of the PCR+/toxin+ class (sensitivity: 0.82 [0.79, 0.84]; specificity: 0.91 [0.90, 0.93]; PPV: 0.82 [0.80, 0.84]. High sensitivity indicates that a negative result could be interpreted as ruling out CDI with high confidence; high specificity indicates that a positive result could be interpreted as ruling in CDI with high confidence.

**Figure 2.**
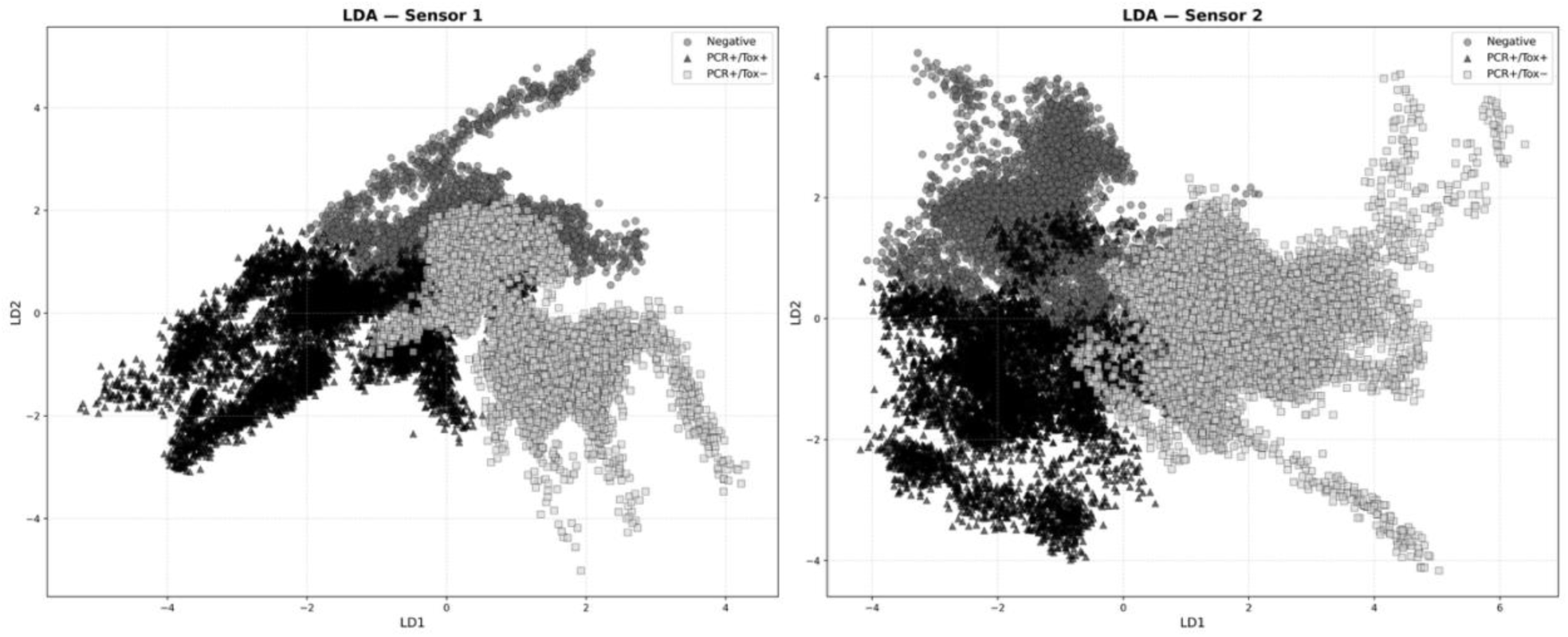
Linear discriminant analysis plot of *C. difficile* class separability in a clinical laboratory setting with Sensors 1 and 2.

**Table 1.** Breakdown of testing and training data split for Sensors 1 and 2.

| Class | Total Observations (n) | Training set (n) | Testing set (n) |
| --- | --- | --- | --- |
| Sensor 1 |  |  |  |
| PCR+/toxin<sup>+</sup><sup>a</sup> | 6,893 | 5,805 (84) | 1,088 (16) |
| PCR+/toxin<sup>-</sup><sup>b</sup> | 8,877 | 7,540 (85) | 1,337 (15) |
| PCR<sup>-</sup><sup>c</sup> | 4,561 | 3,621 (79) | 940 (21) |
| Total | 20,331 | 16,966 (83) | 3,365 (17) |
| Sensor 2 |  |  |  |
| Class | Total Observations (n) | Training set (n) | Testing set (n) |
| PCR+/toxin<sup>+</sup><sup>a</sup> | 7,124 | 5,984 (84) | 1,140 (16) |
| PCR+/toxin<sup>-</sup><sup>b</sup> | 9,992 | 8,674 (87) | 1,318 (13) |
| PCR<sup>-</sup><sup>c</sup> | 6,422 | 5,219 (81) | 1,203 (19) |
| Total | 23,538 | 19,877 (84) | 3,661 (16) |

<sup>a</sup> Stool specimens characterized as polymerase chain reaction positive for *C. difficile* and enzyme immunoassay positive for cytotoxin
<sup>b</sup> Stool specimens characterized as polymerase chain reaction positive for *C. difficile* and enzyme immunoassay negative for cytotoxin
<sup>c</sup> Stool specimens characterized as polymerase chain reaction negative for *C. difficile*

Specificity was also high for the PCR+/toxin– and PCR– classes (0.88 [0.86, 0.90]; and 0.91 [0.89, 0.93], respectively), and sensitivity of the PCR+/toxin– class was high (0.88 [0.86, 0.89]). High specificity for the PCR– class indicates that the presence of a PCR– result would rule out the presence of *C. difficile* with high confidence. Classification performance metrics are further detailed in Table 2. As is shown by the testing data confusion matrix in Figure 3, the greatest amount of misclassification occurred between the PCR+/toxin– and PCR– classes.

**Figure 3.**
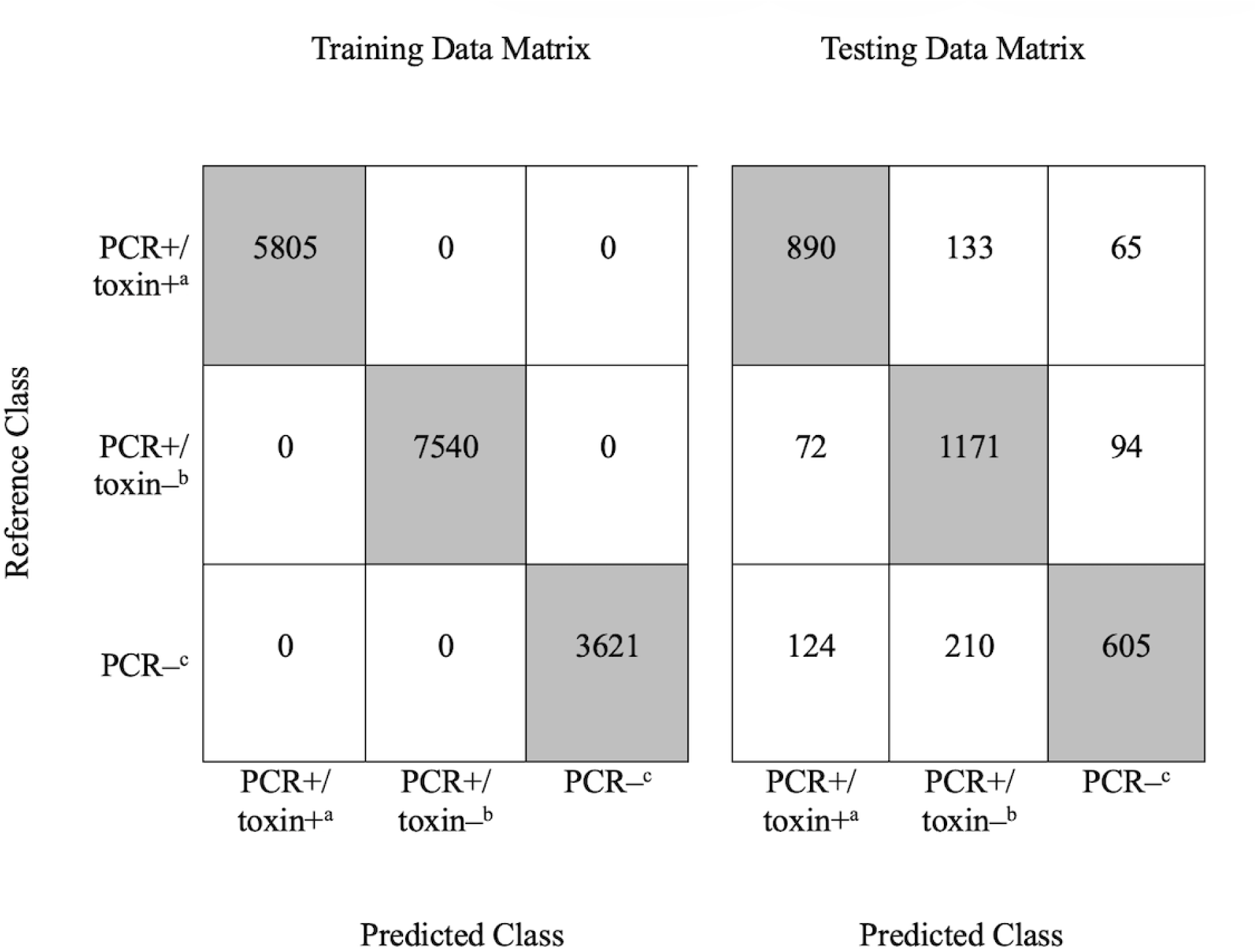
Sensor 1 training and testing confusion matrices. The shaded diagonals represent true positive predictions. Unshaded cells along each reference class row can be interpreted as false negatives with respect to the reference. Unshaded cells along each predicted class column can be interpreted as false positives with reference to the respective reference class.

**Table 2.**
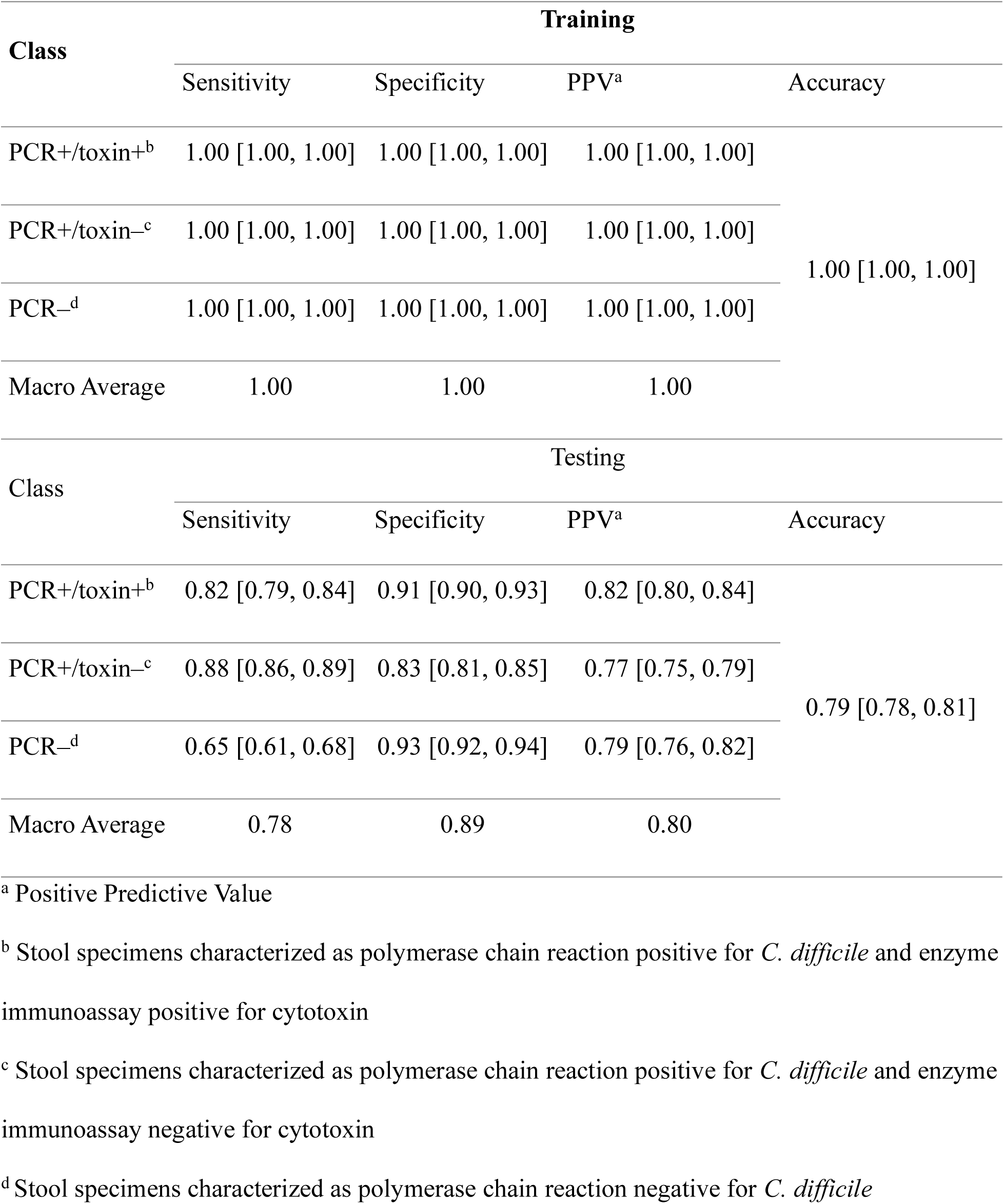
Sensor 1 classification performance metrics for training and testing data sets.

### Sensor 2

A total of 23,538 (7,124 PCR+/toxin+, 9,992 PCR+/toxin–, and 6,422 PCR–) 5-second exposures were captured by Sensor 2. These were split into training and testing datasets further detailed in Table 1. LDA plots revealed that each class formed clusters with some overlap between classes, indicating partial class separability (Figure 2). For testing data, overall accuracy was 0.84 (95% CI: 0.83, 0.85) with high discrimination of the PCR+/toxin+ class (sensitivity: 0.94 [0.92, 0.95]; specificity: 0.93 [0.92, 0.94]; PPV: 0.86 [0.83, 0.87]). High sensitivity indicates that a negative result could be interpreted as ruling out CDI with high confidence; high specificity indicates that a positive result could be interpreted as ruling in CDI with high confidence.

Specificity was also high for the PCR+/toxin– and PCR– classes (0.96 [0.95, 0.96] and 0.88 [0.86, 0.89], respectively), and sensitivity was high for PCR– (0.93 [0.92, 0.95]). High specificity for the PCR– class indicates that the presence of a PCR– result would rule out the presence of *C. difficile* with high confidence. Classification performance metrics are further detailed in Table 3. As is shown by the testing data confusion matrix in Figure 4, the greatest amount of misclassification occurred between the PCR+/toxin– and PCR– classes.

**Figure 4.**
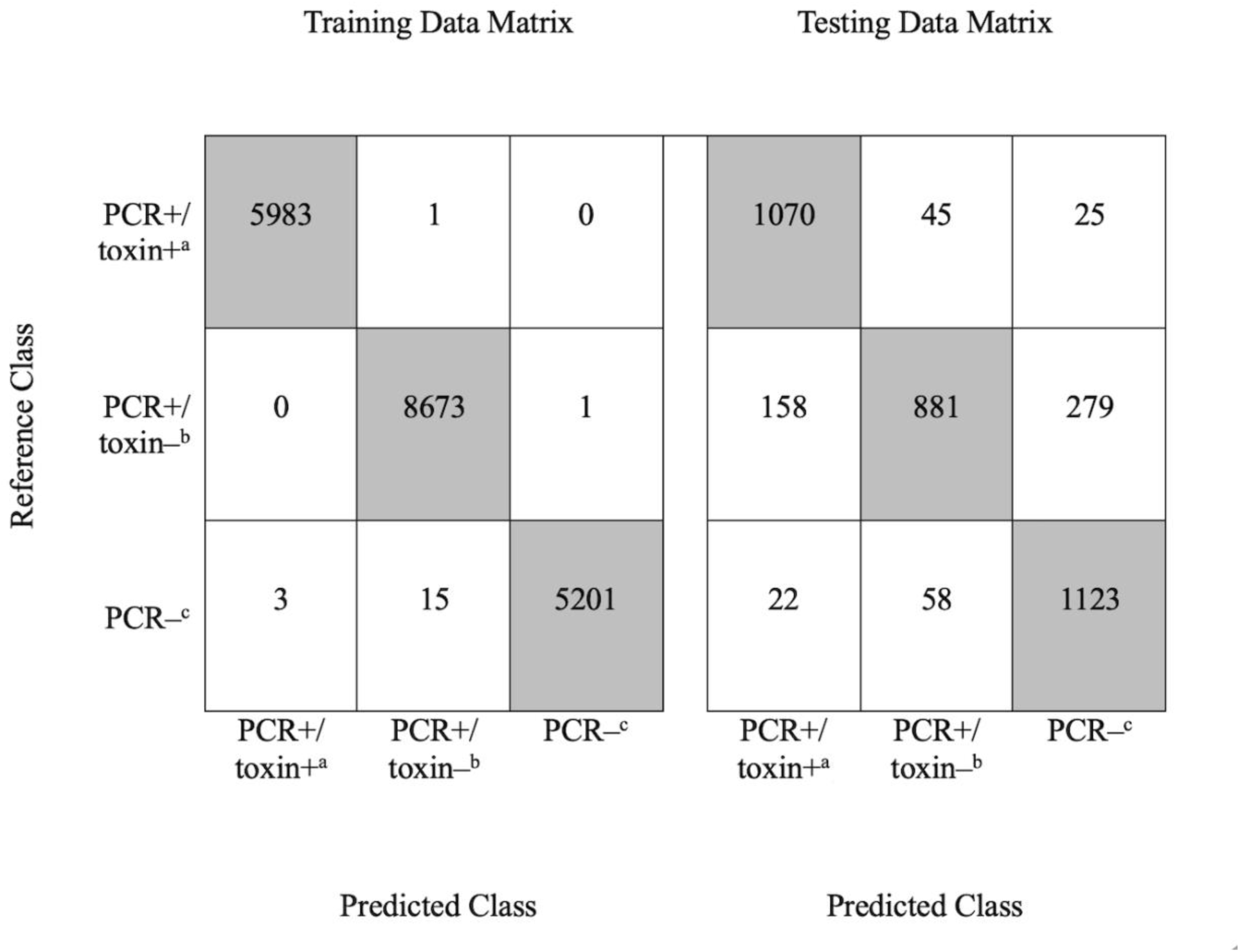
Sensor 2 training and testing confusion matrices. The shaded diagonals represent true positive predictions. Unshaded cells along each reference class row can be interpreted as false negatives with respect to the reference. Unshaded cells along each predicted class column can be interpreted as false positives with reference to the respective reference class.

**Table 3.**
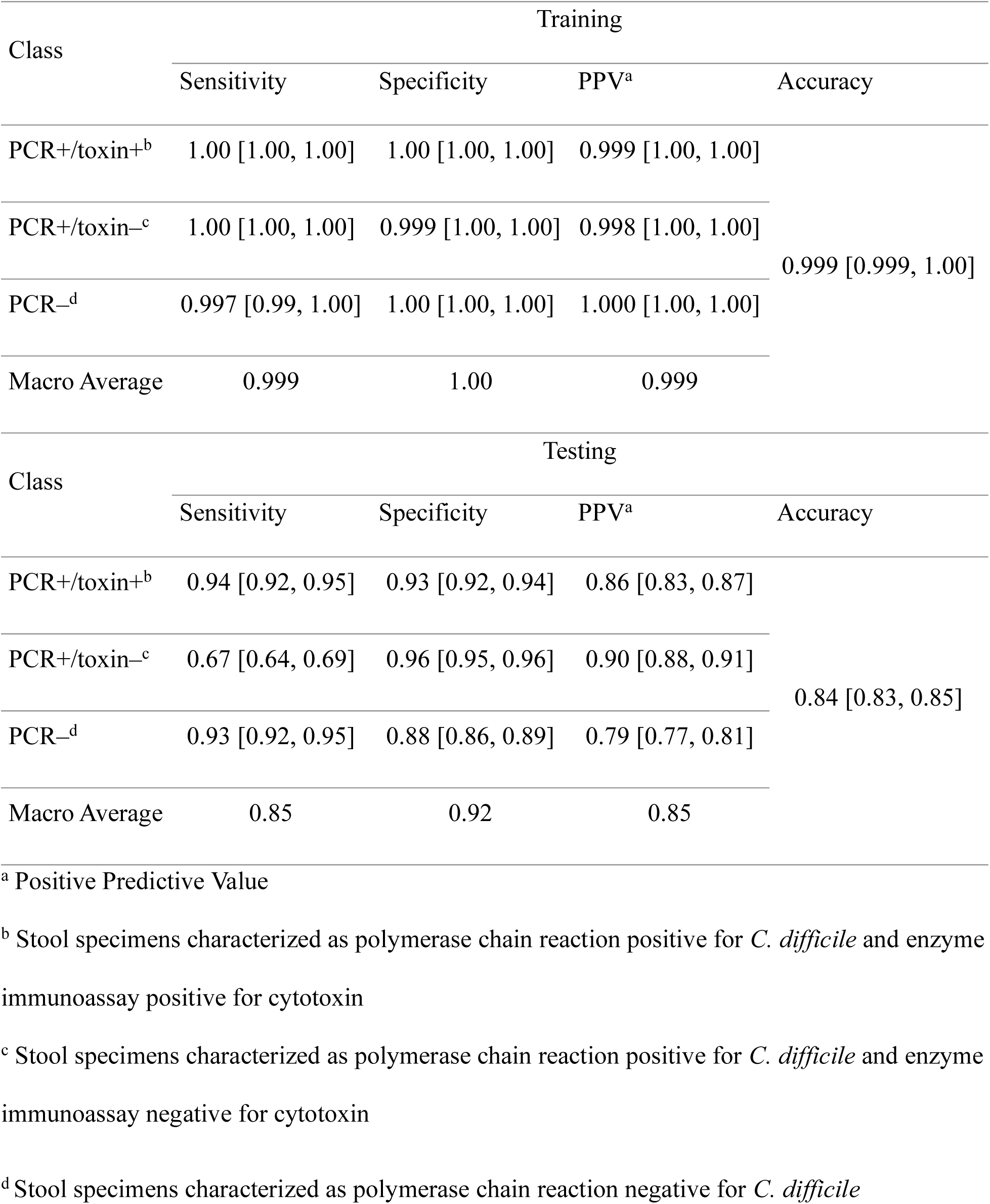
Sensor 2 classification performance metrics for training and testing data sets.

## Discussion

In this pre-screening tool validation study, a multichannel e-nose sensor array differentiated PCR and toxin EIA-characterized *C. difficile* exposure classes under clinical laboratory conditions.

Performance was strongest for PCR+/toxin+ across both Sensors 1 and 2, with high sensitivity (0.82 and 0.94), specificity (0.91 and 0.93), and PPV (0.82 and 0.86), and overall accuracy was also moderate-to-high for both sensors (0.79, 0.84, for Sensors 1 and 2, respectively). These findings are similar to a study that also evaluated an e-nose device’s performance in detecting *C. difficile*, which found an accuracy of 0.84, sensitivity of 0.80, and specificity of 0.85, albeit in a two-class model, unlike the current studies three-class model.^8^ Additionally, the performance of Sensor 2 in classifying PCR+/toxin+ has comparable sensitivity to NAATs and similar specificity to two-test algorithms.^5^ Detection of PCR+/toxin– and PCR– exposures was less consistent between sensors, with moderate sensitivity of PCR+/toxin– in Sensor 2 and moderate sensitivity of PCR– in Sensor 1 (0.67 and 0.65, respectively). Specificity, however, was high in these two classes across devices (Table 1, Table 2).

This study has a few strengths of note. First, three-class model has two distinct advantages in comparison to two-class models. First, this model has the potential utility of identifying PCR+/toxin+, which might indicate an active infection in comparison to PCR+/toxin– and PCR– classes. In practice, a PCR+/toxin+ result would allow for prompt isolation and testing of likely CDI cases. Second, there is a possible biological interpretation to the strong discrimination of PCR+/toxin+: differences in bacterial burden, toxin expression, and/or metabolic activity could influence VOC production, which makes differentiation possible. These differences could also reflect disease progression and severity, and further research could elucidate the connection between VOC profile and clinical outcomes.

The inclusion of Sensor 1 and Sensor 2 in the analysis also serves as a study strength. Differences between the sensors is clearest in the sensitivity of the PCR+/toxin– and PCR– toxin classes, which might reflect intrinsic sensor variability, environmental influences, or channel-specific signal sensitivity. Inclusion of one sensor or the other might have underestimated the potential diagnostic performance of the e-nose device, and the inclusion of both sensors directs future development towards increased reproducibility across devices via further sensor carbon nanotube development, larger validation cohorts and a more advanced 64-channel sensor e-nose.

This study also has a few limitations of note. First, performance was evaluated at the exposure level rather than the specimen level. This approach allowed assessment of sensor response stability and intra-specimen consistency, which are essential in early-stage sensor validation.

However, these data were clustered and not statistically independent because multiple exposures were derived from individual specimens. Confidence intervals presented above are therefore descriptive and may underestimate variance due to within-specimen correlation. Second, this is a single-center study with a moderate sample size of specimens. Future multi-site studies with larger sample sizes analyzed at the specimen level will be required to establish patient-level performance and generalizability. Third, since this study evaluated stool headspace under laboratory containment conditions and was not designed to assess environmental air detection or bedside deployment, these findings are limited to this laboratory environment. Next steps should address this limitation by evaluation of the e-nose in patient care environments, which will include sensing at a greater distance, accounting for ambient VOC interference, and integration into the clinical environment.

Lastly, there was sample quality variation in the stool specimens included in this study. Some samples were stored in preservative whereas others were not. Without preservative, toxin levels decay over time, which might have resulted in PCR+/toxin+ specimens being misclassified as PCR+/toxin– by the e-nose. Given this limitation, the e-nose performed remarkably well in differentiating PCR+/toxin+ exposures from PCR+/toxin– exposures.

Altogether, the results of this study suggest that this e-nose can consistently rule in PCR+/toxin+ stool specimens as CDI cases and rule out PCR– stool specimens as CDI-negative. This study is the first step in validating a continuous environmental detection device for *C. difficile* infection; the next step is validating this device at the bedside. If the results of this study can be replicated in the patient setting there is an opportunity for considerable reduction in CDI cases as well as resource and cost savings in both true-positive CDI cases and true-negative suspected cases.

For true positive CDI cases, earlier detection would spur more timely treatment resulting in less severe symptoms, fewer days in isolation, and reduced lengths of stay. It would also reduce spread within a hospital, and subsequent CDI cases. The resource and cost savings would be considerable, as up to 50% of CDI cases are preventable through early detection.^11^ Most of these resource and cost savings would be attributable to reduced lengths of stay and reduced days in isolation, which make up the bulk of the additional associated costs in CDI patients— conservatively estimated at a per-patient cost of $9400 in inflation-adjusted US dollars.^12,13^ Resource and cost saving would also extend to suspected CDI cases, which are isolated with contact precautions as if they have CDI while waiting for testing. True-negative suspected CDI cases vastly outnumber true-positive suspected cases, so ruling out these cases earlier present significant opportunity cost savings.^14^

### Conclusion

A 16-channel electronic nose sensor array differentiated PCR-characterized CDI exposure classes under clinical laboratory conditions at a VA medical center. Exposure-level analysis demonstrated consistent model discrimination, particularly for toxin-positive specimens. Further prospective validation at the hospital bedside is warranted.

## Data Availability

All data produced in the present study are available upon reasonable request to the authors

## Acknowledgments

The authors would like to thank Lam Nguyen for research coordination, Layla Jones for contributions in the lab.

## Manuscript preparation

Simpli-Fi Automation provided assistance with the study design, data acquisition, and preparing and editing the manuscript.

---

**Table S-1.** Sensor 1 machine learning classification model comparisons.

| Model | Class | Training |  |  | Testing |  |  |
| --- | --- | --- | --- | --- | --- | --- | --- |
|  |  | Sensitivity | Specificity | PPV | Sensitivity | Specificity | PPV |
| Logistic Regression | Negative | 0.668 [0.65, 0.68] | 0.854 [0.85, 0.86] | 0.555 [0.54, 0.57] | 0.890 [0.87, 0.91] | 0.927 [0.92, 0.94] | 0.826 [0.80, 0.85] |
|  | PCR+/Tox+ | 0.698 [0.69, 0.71] | 0.781 [0.77, 0.79] | 0.624 [0.61, 0.64] | 0.796 [0.77, 0.82] | 0.853 [0.84, 0.87] | 0.722 [0.70, 0.75] |
|  | PCR+/Tox− | 0.651 [0.64, 0.66] | 0.873 [0.87, 0.88] | 0.803 [0.79, 0.81] | 0.696 [0.67, 0.72] | 0.891 [0.88, 0.90] | 0.808 [0.78, 0.83] |
|  | Macro average | 0.672 | 0.836 | 0.661 | 0.794 | 0.890 | 0.785 |
| Decision Tree | Negative | 0.994 [0.99, 1.00] | 0.995 [0.99, 1.00] | 0.982 [0.98, 0.99] | 0.588 [0.56, 0.62] | 0.889 [0.88, 0.90] | 0.672 [0.64, 0.70] |
|  | PCR+/Tox+ | 0.987 [0.98, 0.99] | 0.995 [0.99, 1.00] | 0.990 [0.99, 0.99] | 0.675 [0.65, 0.70] | 0.820 [0.80, 0.83] | 0.642 [0.61, 0.67] |
|  | PCR+/Tox− | 0.991 [0.99, 0.99] | 0.995 [0.99, 1.00] | 0.994 [0.99, 1.00] | 0.737 [0.71, 0.76] | 0.796 [0.78, 0.81] | 0.705 [0.68, 0.73] |
|  | Macro average | 0.991 | 0.995 | 0.989 | 0.667 | 0.835 | 0.673 |
| Random Forest | Negative | 1.000 [1.00, 1.00] | 1.000 [1.00, 1.00] | 1.000 [1.00, 1.00] | 0.645 [0.61, 0.68] | 0.934 [0.92, 0.94] | 0.792 [0.76, 0.82] |
|  | PCR+/Tox+ | 1.000 [1.00, 1.00] | 1.000 [1.00, 1.00] | 1.000 [1.00, 1.00] | 0.818 [0.79, 0.84] | 0.914 [0.90, 0.93] | 0.820 [0.80, 0.84] |
|  | PCR+/Tox− | 1.000 [1.00, 1.00] | 1.000 [1.00, 1.00] | 1.000 [1.00, 1.00] | 0.876 [0.86, 0.89] | 0.831 [0.81, 0.85] | 0.773 [0.75, 0.79] |
|  | Macro average | 1.000 | 1.000 | 1.000 | 0.780 | 0.893 | 0.795 |
| XGBoost | Negative | 0.996 [0.99, 1.00] | 1.000 [1.00, 1.00] | 1.000 [1.00, 1.00] | 0.548 [0.52, 0.58] | 0.946 [0.94, 0.95] | 0.797 [0.76, 0.83] |
|  | PCR+/Tox+ | 0.999 [1.00, 1.00] | 1.000 [1.00, 1.00] | 1.000 [1.00, 1.00] | 0.773 [0.75, 0.80] | 0.859 [0.84, 0.87] | 0.724 [0.70, 0.75] |
|  | PCR+/Tox− | 1.000 [1.00, 1.00] | 0.998 [1.00, 1.00] | 0.998 [1.00, 1.00] | 0.912 [0.90, 0.93] | 0.833 [0.82, 0.85] | 0.783 [0.76, 0.80] |
|  | Macro average | 0.998 | 0.999 | 0.999 | 0.744 | 0.879 | 0.768 |
|  |  | Sensitivity | Specificity | PPV | Sensitivity | Specificity | PPV |
| KNN | Negative | 0.994 [0.99, 1.00] | 0.997 [0.99, 1.00] | 0.988 [0.98, 0.99] | 0.809 [0.78, 0.83] | 0.970 [0.96, 0.98] | 0.912 [0.89, 0.93] |
|  | PCR+/Tox+ | 0.985 [0.98, 0.99] | 0.996 [0.99, 1.00] | 0.992 [0.99, 0.99] | 0.642 [0.61, 0.67] | 0.973 [0.97, 0.98] | 0.920 [0.90, 0.94] |
|  | PCR+/Tox− | 0.993 [0.99, 0.99] | 0.993 [0.99, 0.99] | 0.991 [0.99, 0.99] | 0.990 [0.98, 0.99] | 0.779 [0.76, 0.80] | 0.747 [0.73, 0.77] |
|  | Macro average | 0.991 | 0.995 | 0.990 | 0.814 | 0.907 | 0.860 |
| Extra Trees | Negative | 0.994 [0.99, 1.00] | 1.000 [1.00, 1.00] | 1.000 [1.00, 1.00] | 0.657 [0.63, 0.69] | 0.954 [0.94, 0.96] | 0.848 [0.82, 0.87] |
|  | PCR+/Tox+ | 0.995 [0.99, 1.00] | 1.000 [1.00, 1.00] | 1.000 [1.00, 1.00] | 0.686 [0.66, 0.71] | 0.951 [0.94, 0.96] | 0.870 [0.85, 0.89] |
|  | PCR+/Tox− | 1.000 [1.00, 1.00] | 0.994 [0.99, 1.00] | 0.993 [0.99, 0.99] | 0.930 [0.92, 0.94] | 0.736 [0.72, 0.75] | 0.699 [0.68, 0.72] |
|  | Macro average | 0.996 | 0.998 | 0.998 | 0.758 | 0.880 | 0.806 |
| SVM | Negative | 0.815 [0.80, 0.83] | 0.982 [0.98, 0.98] | 0.926 [0.92, 0.94] | 0.754 [0.73, 0.78] | 1.000 [1.00, 1.00] | 1.000 [0.99, 1.00] |
|  | PCR+/Tox+ | 0.814 [0.80, 0.82] | 0.917 [0.91, 0.92] | 0.836 [0.83, 0.84] | 0.692 [0.66, 0.72] | 0.978 [0.97, 0.98] | 0.938 [0.92, 0.95] |
|  | PCR+/Tox− | 0.943 [0.94, 0.95] | 0.893 [0.89, 0.90] | 0.875 [0.87, 0.88] | 0.993 [0.99, 1.00] | 0.741 [0.72, 0.76] | 0.717 [0.70, 0.74] |
|  | Macro average | 0.857 | 0.931 | 0.879 | 0.813 | 0.906 | 0.885 |

**Table S-2.** Sensor 2 machine learning classification model comparisons.

| Model | Class | Training |  |  | Testing |  |  |
| --- | --- | --- | --- | --- | --- | --- | --- |
|  |  | Sensitivity | Specificity | PPV | Sensitivity | Specificity | PPV |
| Logistic Regression | Negative | 0.606 [0.59, 0.62] | 0.924 [0.92, 0.93] | 0.740 [0.73, 0.75] | 0.596 [0.57, 0.62] | 0.879 [0.86, 0.89] | 0.706 [0.68, 0.73] |
|  | PCR+/Tox+ | 0.707 [0.69, 0.72] | 0.742 [0.73, 0.75] | 0.541 [0.53, 0.55] | 0.750 [0.72, 0.77] | 0.655 [0.64, 0.67] | 0.495 [0.47, 0.52] |
|  | PCR+/Tox− | 0.695 [0.69, 0.70] | 0.843 [0.84, 0.85] | 0.774 [0.77, 0.78] | 0.434 [0.41, 0.46] | 0.851 [0.84, 0.86] | 0.622 [0.59, 0.65] |
|  | Macro average | 0.669 | 0.836 | 0.685 | 0.593 | 0.795 | 0.608 |
| Decision Tree | Negative | 0.422 [0.41, 0.43] | 0.998 [1.00, 1.00] | 0.990 [0.98, 0.99] | 0.244 [0.22, 0.27] | 0.953 [0.94, 0.96] | 0.718 [0.67, 0.76] |
|  | PCR+/Tox+ | 0.602 [0.59, 0.61] | 0.846 [0.84, 0.85] | 0.628 [0.62, 0.64] | 0.664 [0.64, 0.69] | 0.731 [0.71, 0.75] | 0.528 [0.50, 0.55] |
|  | PCR+/Tox− | 0.898 [0.89, 0.90] | 0.632 [0.62, 0.64] | 0.654 [0.65, 0.66] | 0.602 [0.58, 0.63] | 0.563 [0.54, 0.58] | 0.437 [0.41, 0.46] |
|  | Macro average | 0.641 | 0.825 | 0.757 | 0.503 | 0.749 | 0.561 |
| Random Forest | Negative | 0.997 [0.99, 1.00] | 1.000 [1.00, 1.00] | 1.000 [1.00, 1.00] | 0.933 [0.92, 0.95] | 0.876 [0.86, 0.89] | 0.787 [0.77, 0.81] |
|  | PCR+/Tox+ | 1.000 [1.00, 1.00] | 1.000 [1.00, 1.00] | 0.999 [1.00, 1.00] | 0.939 [0.92, 0.95] | 0.929 [0.92, 0.94] | 0.856 [0.83, 0.87] |
|  | PCR+/Tox− | 1.000 [1.00, 1.00] | 0.999 [1.00, 1.00] | 0.998 [1.00, 1.00] | 0.668 [0.64, 0.69] | 0.956 [0.95, 0.96] | 0.895 [0.88, 0.91] |
|  | Macro average | 0.999 | 1.000 | 0.999 | 0.847 | 0.920 | 0.846 |
| XGBoost | Negative | 0.669 [0.66, 0.68] | 0.985 [0.98, 0.99] | 0.940 [0.93, 0.95] | 0.606 [0.58, 0.63] | 0.900 [0.89, 0.91] | 0.748 [0.72, 0.77] |
|  | PCR+/Tox+ | 0.693 [0.68, 0.70] | 0.929 [0.93, 0.93] | 0.809 [0.80, 0.82] | 0.671 [0.64, 0.70] | 0.830 [0.81, 0.84] | 0.641 [0.61, 0.67] |
|  | PCR+/Tox− | 0.953 [0.95, 0.96] | 0.753 [0.74, 0.76] | 0.749 [0.74, 0.76] | 0.618 [0.59, 0.64] | 0.711 [0.69, 0.73] | 0.546 [0.52, 0.57] |
|  | Macro average | 0.772 | 0.889 | 0.833 | 0.632 | 0.814 | 0.645 |
|  |  | Sensitivity | Specificity | PPV | Sensitivity | Specificity | PPV |
| KNN | Negative | 0.966 [0.96, 0.97] | 0.996 [0.99, 1.00] | 0.988 [0.98, 0.99] | 0.874 [0.85, 0.89] | 0.891 [0.88, 0.90] | 0.797 [0.77, 0.82] |
|  | PCR+/Tox+ | 0.981 [0.98, 0.98] | 0.992 [0.99, 0.99] | 0.982 [0.98, 0.98] | 0.959 [0.95, 0.97] | 0.840 [0.82, 0.85] | 0.731 [0.71, 0.75] |
|  | PCR+/Tox− | 0.991 [0.99, 0.99] | 0.982 [0.98, 0.98] | 0.977 [0.97, 0.98] | 0.594 [0.57, 0.62] | 0.973 [0.97, 0.98] | 0.926 [0.91, 0.94] |
|  | Macro average | 0.979 | 0.990 | 0.982 | 0.809 | 0.901 | 0.818 |
| Extra Trees | Negative | 1.000 [1.00, 1.00] | 1.000 [1.00, 1.00] | 1.000 [1.00, 1.00] | 0.889 [0.87, 0.91] | 0.907 [0.90, 0.92] | 0.824 [0.80, 0.84] |
|  | PCR+/Tox+ | 1.000 [1.00, 1.00] | 1.000 [1.00, 1.00] | 1.000 [1.00, 1.00] | 0.932 [0.92, 0.94] | 0.861 [0.85, 0.87] | 0.752 [0.73, 0.77] |
|  | PCR+/Tox− | 1.000 [1.00, 1.00] | 1.000 [1.00, 1.00] | 1.000 [1.00, 1.00] | 0.641 [0.61, 0.67] | 0.954 [0.94, 0.96] | 0.888 [0.87, 0.91] |
|  | Macro average | 1.000 | 1.000 | 1.000 | 0.821 | 0.907 | 0.821 |
| SVM | Negative | 0.862 [0.85, 0.87] | 0.996 [0.99, 1.00] | 0.987 [0.98, 0.99] | 0.818 [0.80, 0.84] | 0.913 [0.90, 0.92] | 0.820 [0.80, 0.84] |
|  | PCR+/Tox+ | 0.870 [0.86, 0.88] | 0.961 [0.96, 0.96] | 0.907 [0.90, 0.91] | 0.868 [0.85, 0.89] | 0.904 [0.89, 0.92] | 0.805 [0.78, 0.83] |
|  | PCR+/Tox− | 0.967 [0.96, 0.97] | 0.894 [0.89, 0.90] | 0.876 [0.87, 0.88] | 0.763 [0.74, 0.79] | 0.903 [0.89, 0.92] | 0.817 [0.80, 0.84] |
|  | Macro average | 0.900 | 0.950 | 0.923 | 0.816 | 0.907 | 0.814 |

